# Evaluation of a rapid lateral flow calprotectin test for the diagnosis of prosthetic joint infection

**DOI:** 10.1101/19004473

**Authors:** Alexander J. Trotter, Rachael Dean, Celia E. Whitehouse, Jarle Mikalsen, Claire Hill, Roxanne Brunton-Sim, Gemma L. Kay, Majeed Shakokhani, Alexander Durst, John Wain, Iain McNamara, Justin O’Grady

## Abstract

**Background:** Microbiological diagnosis of prosthetic joint infection (PJI) relies on culture techniques that are slow and insensitive. Rapid tests are urgently required to improve patient management. Calprotectin is a neutrophil biomarker of inflammation that has been demonstrated to be effective for the diagnosis of PJI. A calprotectin based lateral flow test has been developed for the rapid detection of PJI using synovial fluid samples.

**Methods:** A convenience series of 69 synovial fluid samples from patients at the Norfolk and Norwich University Hospitals (NNUH) were collected intraoperatively from 52 hip and 17 knee revision operations. Calprotectin levels were measured using a new commercially available lateral flow assay for PJI diagnosis (Lyfstone). For all samples, synovial fluid was pipetted onto the lateral flow device and the signal was read using a mobile phone app after 15 minutes incubation at room temperature.

**Results:** According to the Musculoskeletal Infection Society (MSIS) criteria, 24 patients were defined as PJI positive and the remaining 45 were negative. The overall accuracy of the lateral flow test against the MSIS criteria was 75%. The test had a sensitivity and specificity of 75% and 76% respectively with a positive predictive value (PPV) of 62% and a negative predictive value (NPV) of 85%. Discordant results were then reviewed by the clinical team using available patient data to develop an alternative gold standard for defining presence/absence of infection (MSIS+). Compared to MSIS+, the test showed an overall accuracy of 83%, sensitivity and specificity of 95% and 78% respectively, a PPV of 62% and an NPV of 98%. Test accuracy for hip revisions was 77% and for knee revisions was 100%.

**Conclusions:** This study demonstrates that the calprotectin lateral flow assay is an effective diagnostic test for PJI. Our data suggests that the test is likely to generate false positive results in patients with metallosis and gross osteolysis.

## Introduction

Each year in the UK, approximately 160,000 primary hip and knee replacements are performed plus an additional 14,000 revision operations. Prosthetic Joint Infection (PJI) is responsible for 14% of these revisions [1].

Diagnosis of PJI remains a challenge and guidelines vary between countries. Low-grade PJIs are particularly difficult to diagnose. They are commonly caused by bacteria which do not have a clear pathogenic role and often contaminate tissue samples, making culture results difficult to interpret [2]. These organisms are also less likely to trigger an increase in inflammatory makers [3]. The inability to differentiate between low-grade infections and aseptic loosening, leads to patients undergoing numerous investigations and unnecessary two-stage revisions. This treatment option comes with a higher cost both to the to the healthcare system and accompanying patient morbidity, along with an associated higher complication rate than one-stage revisions [4]. For this group of patients especially there is a need for a diagnostic test that can reliably exclude PJI.

The Musculoskeletal Infection Society (MSIS) published a list of criteria designed to standardise the diagnosis of PJI [5]. The criteria require a diagnostic work-up combining preoperative and intraoperative findings with selected inflammatory markers and culture results. MSIS criteria are useful but fallible, due to the heavy weighting placed on microbiological culture results, which are known to have sub-optimal specificity and sensitivity.

In 2018, the MSIS criteria were revised due to the emergence of new PJI biomarkers including serum D-dimer, synovial alpha-defensins and synovial leukocyte esterase [6]. These tests can be expensive and their application in clinical practice is currently limited. An alternative biomarker is synovial fluid calprotectin. Calprotectin is a protein complex released during inflammation that makes up 60% of all soluble proteins in neutrophils [7]. Neutrophils are recruited to sites of inflammation and infection response; therefore, high amounts of neutrophils biomarkers are expected to be seen in infected patient samples [8].

Calprotectin is routinely used to screen for inflammatory bowel disease and has been shown to detect relapses in rheumatoid arthritis [9, 10]. In a recent study, a stool calprotectin test was used off label for PJI diagnosis demonstrating a negative predictive value (NPV) of 94.4% [11]. This demonstrates the utility of calprotectin for PJI diagnosis and the need for a validated lateral flow test.

Lyfstone have recently developed a lateral flow calprotectin test for the diagnosis of PJI which has passed the European IVD regulatory approval process (98/79/EC). We conducted the first clinical evaluation of this new technology in a retrospective study on samples from suspected PJI or aseptic loosening cases to assess the sensitivity, specificity, NPV and PPV.

## Materials and Methods

A total of 73 synovial fluid samples collected as a convenience series and stored in the Biorepository at the University of East Anglia between February 2016 and January 2019 as part of routine clinical practice from patients treated at the Norfolk and Norwich University Hospitals (NNUH) were retrospectively tested for calprotectin levels with the Lyfstone lateral flow test. Informed consent was obtained from all patients included in the study. Ethical approval was provided by University of East Anglia, Faculty of Medicine and Health Sciences Research Ethics Committee, project reference: 2016/17 21 SE. Samples were only tested if of sufficient volume (≥100µL) and results for microbiological culture, frozen-section histology and serum C-Reactive Protein (CRP) were available. Four samples were removed from the analysis due to incomplete/erroneous patient data (two had no pre-operative notes and two were from a patient who had two separate surgeries, but the samples had the same date). The final total of 69 samples were included in the dataset (37 male, 32 female, age range 45-89), 52 were taken during hip revision surgery and 17 from knee revisions.

Cases were classified as either infected or aseptic based on the MSIS criteria (Table 1). A case was deemed as infected if the patient presented either with a major criterion, i.e. two or more cultures of the same organism or the presence of a sinus tract; or met three minor criteria i.e. elevated serum CRP levels, purulence, one positive microbiological culture and/or positive histological analysis of periprosthetic tissue for inflammation. Other minor criteria from the 2018 MSIS criteria were not considered as many of those tests are not used as standard in the UK or routine at NNUH. Clinical information was not made available to performers of the index test before testing.

**Table 1.** Scoring based definition for PJI using available tests from the 2018 MSIS criteria.

| Major Criteria (one or more of the following) | Decision |
| --- | --- |
| Two positive cultures of the same organism | Infected |
| Sinus tract with evidence of communication to the joint or visualization of the prosthesis |  |

| Minor Criteria | Score | Decision |
| --- | --- | --- |
| Elevated serum CRP ($\geq 10$ mg/L) | 2 | $\geq 6$ Infected |
| Positive histology | 3 |  |
| Positive purulence | 3 | <6 Aseptic |
| Single positive culture | 2 |  |

Discrepant results were investigated by one of the senior authors (IM - Consultant Orthopaedic Surgeon) and team by reviewing patient medical records, surgical notes and X-rays to make a definite decision on whether the patients were managed by the clinical team as infected (MSIS+).

The Lyfstone calprotectin test was carried out according to the manufacturer’s instructions. Synovial fluid samples (20µl) were diluted in a pre-mixed dilution buffer (2 ml – 101x dilution) and added to the test cassette (80µl). Gold-conjugated antibody complexes then bind calprotectin and travel along the membrane within the cassette and further bind to immobilised calprotectin-specific antibodies to form a visible test line. Any remaining gold-conjugated antibody not bound to calprotectin is immobilised on a control line. After 15 minutes incubation at room temperature the cassette was photo-imaged, and the calprotectin level calculated by the Lyfstone smartphone application (Figure 1). The colour intensity of the test line is proportional to the concentration of calprotectin in the sample. A calprotectin result of ≤14 mg/L was considered a negative result according to the cut-off between low and moderate risk of infection by the Lyfstone smartphone app at the time of testing. Moderate (14-50 mg/L) and severe (>50 mg/L) risk of infection categories were grouped together as positive [12].

**Figure 1.**
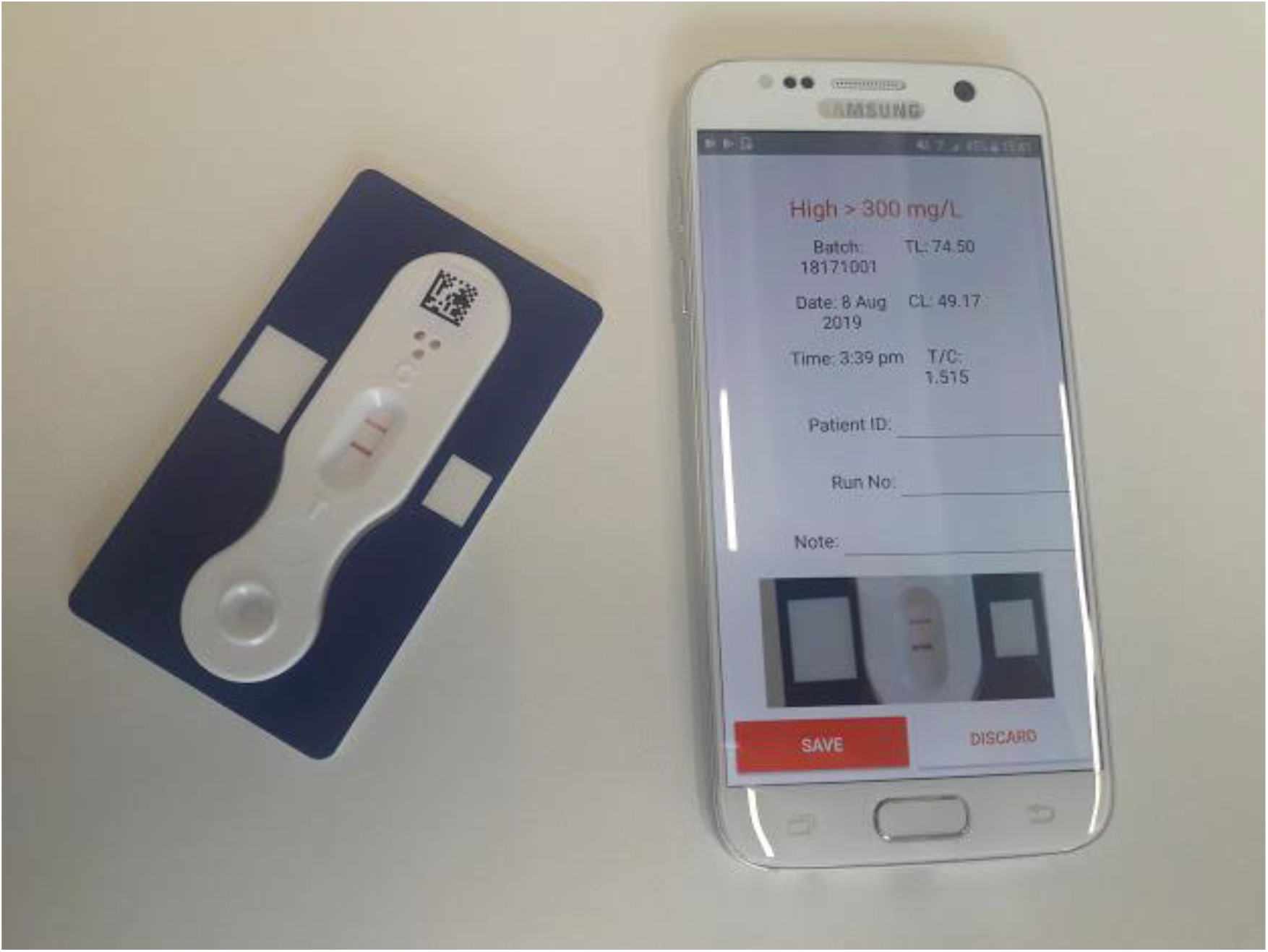
Lyfstone App report (right) from Lyfstone Calprotectin test (left).

Microbiological culture was performed on all tissue and fluid specimens (recommended that 3-5 tissue/fluid samples sent for testing) for 48 hours on blood agar and chocolate agar, five days on fastidious anaerobe agar, and five days in cooked meat broth before 48-hour subculture on fastidious anaerobe agar, chocolate agar, sabouraud agar and in fastidious antimicrobial neutralization bottles according to UK Standards for Microbiology Investigations B 44 [13]. Histology was performed by frozen section microscopy according to UKAS ISO 8405 [14]; Serum CRP was performed according to UKAS ISO 10294 [15].

## Results

### Lyfstone test performance compared to MSIS gold standard

The overall sensitivity and specificity of the calprotectin test for the diagnosis of PJI compared to MSIS gold standard was 75% and 75.6% respectively. The PPV was 62.1%, the NPV 85% and the accuracy was 75.4% (Table 2 and 3).

**Table 2.**
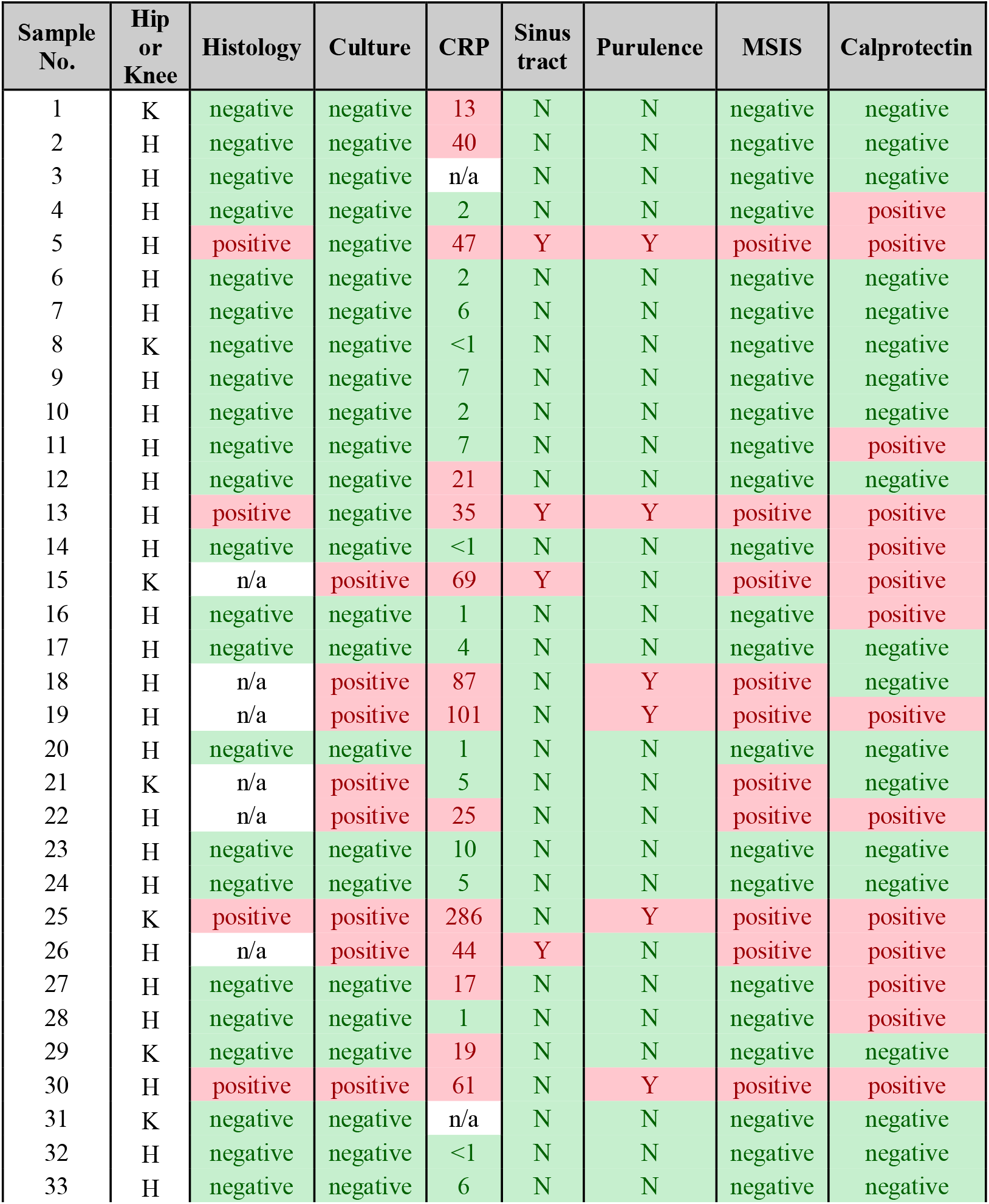
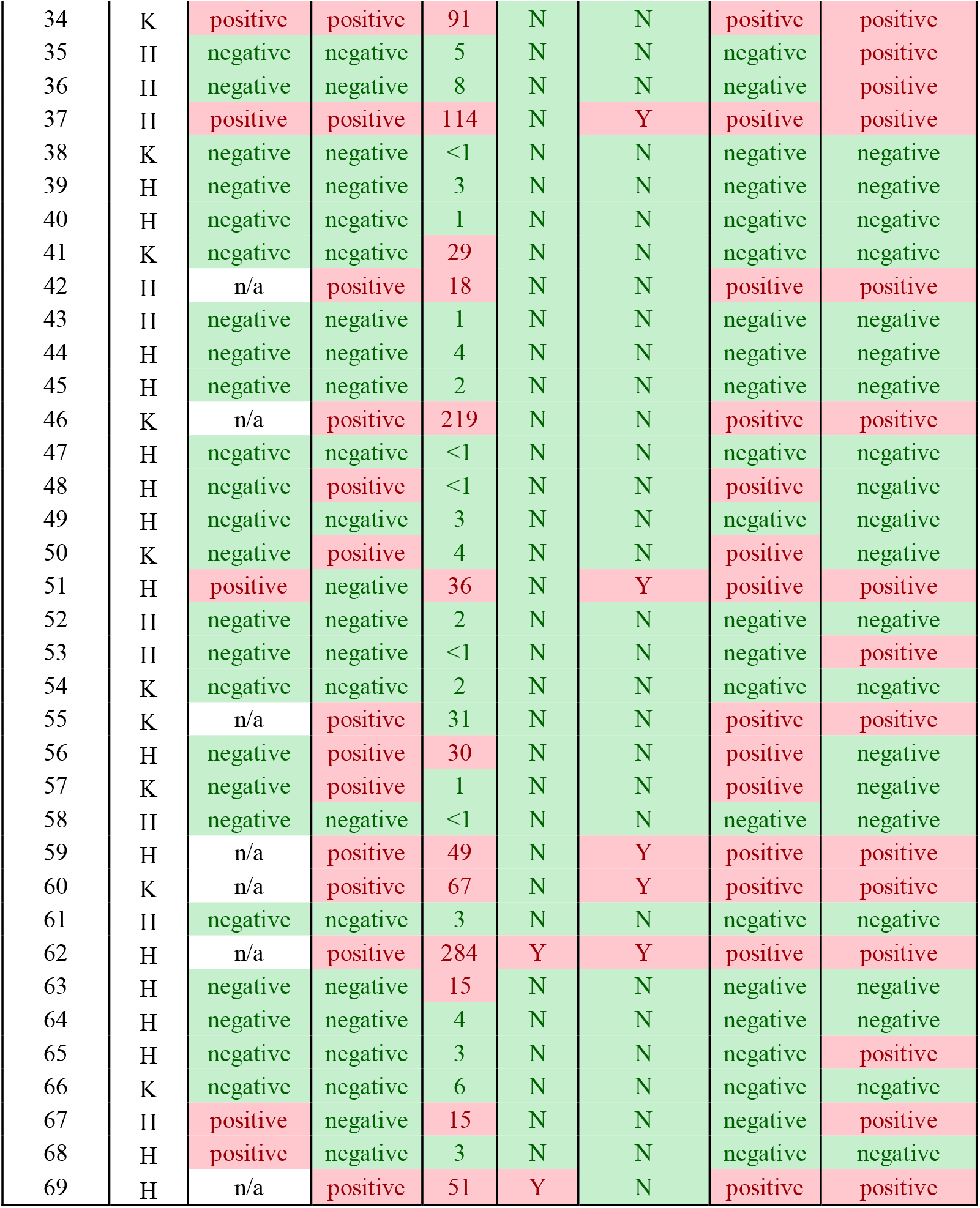
MSIS and calprotectin test results. Histology: positive/negative for signs of inflammation by frozen section microscopy. Culture: positive/negative for two or more cultures of the same organism from periprosthetic tissue and fluid samples. CRP: positive when serum CRP level ≥10 mg/L. Sinus tract: Y = presence of sinus tract. Purulence: Y = visible wound purulence. MSIS: positive/negative according to scoring in table 1. Calprotectin: positive when ≥ 14 mg/L synovial calprotectin measured by Lyfstone calprotectin test.

**Table 3.** Performance of calprotectin test on tested synovial fluid samples. All samples, n=69; Hips, n=52; Knees, n=17

| Lyfstone test | Gold standard |  |  |  |  |  |
| --- | --- | --- | --- | --- | --- | --- |
|  | MSIS |  |  | MSIS+ |  |  |
|  | All samples | Hips | Knees | All samples | Hips | Knees |
| **Sensitivity** | **75.00%** | **80%** | **66.67%** | **94.74%** | **92.31%** | **100%** |
| (95% CI) | (53.29% - 90.23%) | (51.91% - 95.67%) | (29.93% - 92.51%) | (73.97% - 99.87%) | (63.97% - 99.81%) | (54.07% - 100%) |
| **Specificity** | **75.56%** | **70.27%** | **100%** | **78.00%** | **71.79%** | **100%** |
| (95% CI) | (60.46% - 87.12%) | (53.02% - 84.13%) | (63.06% - 100%) | (64.04% - 88.47%) | (55.13% - 85%) | (71.51% - 100%) |
| **PPV** | **62.07%** | **52.17%** | **100%** | **62.07%** | **52.17%** | **100%** |
| (95% CI) | (48.23% - 74.19%) | (38.48% - 65.55%) | (100% - 100%) | (49% - 73.6%) | (39.23% - 64.83%) | (100% - 100%) |
| **NPV** | **85.00%** | **89.66%** | **72.73%** | **97.50%** | **96.55%** | **100%** |
| (95% CI) | (73.54% - 92.04%) | (75.51% - 96.06%) | (51.42% - 87.04%) | (85.2% - 99.62%) | (80.83% - 99.47%) | (100% - 100%) |
| **Accuracy** | **75.36%** | **73.08%** | **82.35%** | **82.61%** | **76.92%** | **100%** |
| (95% CI) | (63.51% - 84.95%) | (58.98% - 84.43%) | (56.57% - 96.2%) | (71.59% - 90.68%) | (63.16% - 87.47%) | (80.49% - 100%) |

A total of 24 cases were classed as infected and 45 cases aseptic according to MSIS criteria. Of the infected cases, 21 were positive by routine microbiology alone (>2 cultures positive cultures for the same organism). Two of the remaining cases were positive by histology, elevated serum CRP and one positive culture (*Collinsella aeofasciens* and ‘Diptheroids’) and the remaining infected case was positive by a combination of histology, CRP and purulence (Table 2).

In this infected group there were 18 true positive and 6 false negative results by calprotectin. The reported organisms in these false negative cases were *Cutibacterium acnes* (n=3), *Bacteroides fragilis* (n=1), *Staphylococcus epidermidis* (n=1), and a polymicrobial infection of *Pseudomonas aeruginosa* with *Corynebacterium tuberculostearicum* (n=1).

The remaining 45 cases were aseptic by MSIS criteria and of these 34 were true negative and 11 were false positive by calprotectin. Of the 11 false positive cases, one case was positive for inflammation by histology and elevated CRP and one was CRP positive without significant histology. The remaining nine false positive samples were negative for any MSIS criteria tested.

In the hip revision surgery group (n=52) the test had a sensitivity of 80% and specificity of 70.3%. The PPV and NPV were 52.2% and 89.7% respectively with an overall test accuracy of 73.1%. All 11 false positive results in the study were hip revisions. In the knee revision surgery group (n=17) the test was 66.7% sensitive and 100% specific. With no false positive results, the PPV was 100% but the NPV was 72.7% (Supplementary table 1). Overall test accuracy was higher than in the hip group at 82.4%.

### Lyfstone test performance compared to MSIS+ gold standard

Overall test accuracy compared to the MSIS+ standard was 82.6%. Sensitivity and specificity were 94.7% and 78% respectively, with PPV of 62.1% and NPV of 97.5% (Table 3).

In the hip group, medical record and radiological review revealed 7/11 false positive results were associated with metallosis from patients who were being revised for metal on metal hip implants, an expected trigger for gross inflammation [16] thereby resulting in high calprotectin and false positive results. The remaining four false positive cases had aseptic loosening listed as the initial indication for operation. Review of pre-operative radiographs revealed gross osteolysis in 3 out of 4 cases, with gross destruction of either proximal femur or acetabulum. This again is likely to cause false positive calprotectin results. One-year follow-up of one of the three patients (patient 35) revealed that they were still experiencing pain in their joint and their CRP had raised from 1 mg/L to 19 mg/L (Supplementary table 1), indicating possible infection, but as yet still under investigation by the treating teams. Removal of metallosis and severe osteolysis cases from the dataset improved the specificity, PPV and overall accuracy of the test both against MSIS and MSIS+ (Table 4).

**Table 4.** Performance of calprotectin test on tested synovial fluid samples, excluding metallosis and severe osteolysis cases. MSIS (excluding metallosis and severe osteolysis cases): calprotectin result against MSIS criteria (n=53) for infection excluding all samples associated with metallosis and severe osteolyisis. MSIS+ (excluding metallosis and severe osteolysis cases): calprotectin result against MSIS criteria (n=53) for infection with discrepant samples investigated by clinical follow-up excluding all samples associated with metallosis and severe osteolyisis.

| Lyfstone test | Gold Standard |  |
| --- | --- | --- |
|  | MSIS<br>(Excluding metallosis and severe osteolysis cases) | MSIS+<br>(Excluding metallosis and severe osteolysis cases) |
| **Sensitivity**<br>(95% CI) | **71.43%**<br>(47.82% - 88.72%) | **93.75%**<br>(69.77% - 99.84%) |
| **Specificity**<br>(95% CI) | **96.88%**<br>(83.78% - 99.92%) | **97.30%**<br>(85.84% - 99.93%) |
| **PPV**<br>(95% CI) | **93.75%**<br>(68.14% - 99.06%) | **93.75%**<br>(68.36% - 99.05%) |
| **NPV**<br>(95% CI) | **83.78%**<br>(72.37% - 91.06%) | **93.70%**<br>(84.36% - 99.59%) |
| **Accuracy**<br>(95% CI) | **86.79%**<br>(74.66% - 94.52%) | **96.23%**<br>(87.02% - 99.54%) |

The three false negative results within the hip subset were all MSIS positive by culture alone (two or more cultures of the same organism). Patient 18 was culture positive for *B. fragilis*, isolated from all cultured tissue samples, along with a high CRP and noted joint purulence. Calprotectin was recorded at 13.36 mg/L which falls just below the threshold value for a positive result (≥14 mg/L) (Supplementary table 1). Sample 48 was culture positive for *C. acnes* but negative by both histology and CRP. The calprotectin test was negative with 0.0 mg/L. The initial indication for operation for this case was aseptic loosening and a single stage revision was performed - the *C. acnes* report was dismissed as contamination and the patient was not treated for infection. One-year follow-up of the patient showed no signs of infection indicating that the calprotectin result correlated with the clinical findings. Sample 56 was a similar case of an aseptic loosening revision yielding a significant culture results (*P. aeruginosa* and Corynebacterium) but no detectable synovial fluid calprotectin. The patient had a single stage revision and upon a two-month follow-up, showed no sign of infection, indicating a true negative calprotectin result. The clinical review (MSIS+) increases the sensitivity of the Lyfstone test from 75% to 94.74% (Table 3).

In the knee group, three false negative results were recorded; two cases of *C. acnes* and one case reporting *S. epidermidis*. Both *C. acnes* cases (Samples 50 and 57) were patients displaying no pre-operative indication of infection and were treated as aseptic loosening. The clinical review found no complications after patient follow up suggesting both cases were aseptic and the calprotectin results were correct. Sample 21 (*S. epidermidis*) was taken during the second stage of revision surgery performed one year after the first stage operation. Clinical review revealed the patient was originally treated for gross infection. At the time of the second stage revision, the patient showed no overt signs of infection, so treatment progressed to implantation of the prosthesis. Post-operative microbiology grew a culture of *S. epidermidis* from 3/6 samples taken at the time of surgery. At the one-year follow-up the patient presented no sign of infection, confirming the calprotectin result to be correct, thus far. Overall, the Lyfstone test was 100% accurate for the diagnosis of PJI in knee samples (n=17 – Tables 2 and 3).

## Discussion

PJI is a serious complication of total joint arthroplasty with associated patient morbidity and the economic burden of treatment. Microbiological culture techniques are slow and insensitive and although new diagnostic tests are being developed, none of the current methods are capable of reliably diagnosing PJI.

Rapid biomarker-based tests have the potential to assist clinicians in making a pre-operative diagnosis. Early differentiation of PJI from aseptic loosening may reduce the number of unnecessary two-stage revisions performed on patients with a presumptive diagnosis of low-grade infection.

Zimmer Biomet have developed the Synovasure® Test, which functions very similarly to the Lyfstone calprotectin test, measuring another neutrophil released antimicrobial peptide, α-defensin. Studies using Synovasure® have shown varying results with sensitivities ranging from 67% to 100% and specificities ranging from 82.4% to 100% against the MSIS criteria [17-25]. A systematic review summarising the results of seven prospective trials found the Synovasure® lateral flow test had an overall sensitivity of 85% and a specificity of 90% [26].

The performance of the Lyfstone calprotectin rapid test was determined using MSIS and MSIS+ standards. Comparing the results to the MSIS criteria, the PPV was 62% and NPV was 85%. Further analysis revealed that the low PPV is heavily affected by the presence of metallosis cases from revision of metal on metal implants within our sample set. Removing these samples yields a PPV of 79%. The use of biomarker based diagnostic tests is known to be unreliable in patients with metal-on-metal implants as it can cause gross inflammation producing false positive results [27]. This effect was observed in our study with seven of 13 metal-on-metal revisions producing false positive results (true positive = 3, true negative = 3). Patients with evidence of severe osteolysis also contributed a disproportionate number of false positive results (n=3). Removing both metallosis and osteolysis cases resulted in a PPV of 93.8%.

Compared to MSIS, six false negative results were observed. Three of these were MSIS positive by culture of *C. acnes*. This organism is a common lab contaminant and not typically considered a cause of infection when isolated from hip and knee tissues [28-30]. In fact, 5/6 false negative Lyfstone test results were shown to be true negatives when compared to the MSIS+ clinical standard, highlighting the need for improved diagnostics. Only one case of infection was missed (case 18) in which the calprotectin result was very close to positive (13.36mg/L, < 1 mg/L below the positive cut-off). If future optimisation of the test sets a lower threshold value for infection, then this case would have been classified as positive. Such a readjustment of the calprotectin cut-off values may improve test performance in detecting low-grade infections.

The sensitivity of the Lyfstone test increased from 75% to 94.74% when using the MSIS+ compared to the MSIS criteria and the NPV increased from 85% to 97.5%. There was only a modest increase in specificity and no increase in PPV. However, excluding both metallosis and osteolysis patients increased specificity to 97.3% and PPV to 93.75%.

Test accuracy was different for hip and knee revisions. False positive results were only observed in hip revisions, not in knee revision as there was no metal-on-metal reactions or significant osteolysis in the knee revision group. Compared to MSIS+, the Lyfstone test was 100% accurate for the diagnosis of PJI in knee revisions, however the sample size is small (n=17) and more data is required to confirm these findings.

One limitation of this study is the use of retrospective samples, however, by testing all eligible samples in the biorepository we believe that we did not introduce any sample selection bias. Another limitation was the samples had been frozen for storage before testing and the freeze-thaw process may result in leukocyte cell lysis and an increase in calprotectin. As the test is validated for use on fresh synovial fluid samples, the recommended cut-offs may not necessarily be appropriate for our samples.

It is worth noting that blood contamination of the samples had no impact on the accuracy of the test results, something the Synovasure® lateral flow test warns may lead to false negative results [31].

In conclusion, the Lyfstone calprotectin assay shows good potential as a rapid diagnostic test for PJI due to its high sensitivity and NPV. The test has a low PPV compared to MSIS and MSIS+ but the false positive results are mostly in patients with metal on metal reactions or significant osteolysis, which could serve as contraindications for use. Prospective studies on fresh samples in appropriate patient populations are required to accurately define test performance.

## Data Availability

All data generated or analysed during this study are included in this published article (and its supplementary information files).

## Author contributions

Research design – AT, JM, IM, JOG; Sample testing – AT, RD, RBS; Clinical data acquisition – RD, CW, RBS; Identification, consent and collection of samples – CW, CH, MS, AD; Patient reviews – IM; Manuscript writing – AT, RD, GK, IM, JOG; Training and support – JM; Clinical management and setup of trial – IM, JW; Manuscript review – all authors

